# Development and comparison of natural language processing models for abdominal aortic aneurysm repair identification and classification using unstructured electronic health records

**DOI:** 10.1101/2024.12.11.24318852

**Authors:** Daniel Thompson, Reza Mofidi

**Author notes:** Corresponding author: Daniel Thompson.

## Abstract

**Background:** Patient identification for national registries often relies upon clinician recognition of cases or retrospective searches using potentially inaccurate clinical codes, potentially leading to incomplete data capture and inefficiencies. Natural Language Processing (NLP) offers a promising solution by automating analysis of electronic health records (EHRs). This study aimed to develop NLP models for identifying and classifying abdominal aortic aneurysm (AAA) repairs from unstructured EHRs, demonstrating proof-of-concept for automated patient identification in registries like the National Vascular Registry.

**Method:** Using the MIMIC-IV-Note dataset, a multi-tiered approach was developed to identify vascular patients (Task 1), AAA repairs (Task 2), and classify repairs as primary or revision (Task 3). Four NLP models were trained and evaluated using 4,870 annotated records: scispaCy, BERT-base, Bio-clinicalBERT, and a scispaCy/Bio-clinicalBERT ensemble. Models were compared using accuracy, precision, recall, F1-score, and area under the receiver operating characteristic curve.

**Results:** The scispaCy model demonstrated the fastest training (2 mins/epoch) and inference times (2.87 samples/sec). For Task 1, scispaCy and ensemble models achieved the highest accuracy (0.97). In Task 2, all models performed exceptionally well, with ensemble, scispaCy, and Bio-clinicalBERT models achieving 0.99 accuracy and 1.00 AUC. For Task 3, Bio-clinicalBERT and the ensemble model achieved an AUC of 1.00, with Bio-clinicalBERT displaying the best overall accuracy (0.98).

**Conclusion:** This study demonstrates that NLP models can accurately identify and classify AAA repair cases from unstructured EHRs, suggesting significant potential for automating patient identification in vascular surgery and other medical registries, reducing administrative burden and improving data capture for audit and research.

## Introduction

The integration of Natural Language Processing (NLP) in vascular surgery has gained increasing interest, due to its ability to help analyse data from electronic health records (EHRs), provide feedback on current practice and develop support systems to optimise care [1]. NLP techniques have evolved significantly over the years. Initially, text classification tasks relied on rule-based approaches, which, despite being somewhat effective, faced limitations in managing the complexity and variability of natural language, particularly in the biomedical field [2]. These techniques have since evolved from simple heuristic-based models to sophisticated neural network-based architectures [3]. The integration of word embeddings, contextualised embeddings, and transfer learning has significantly enhanced the capability of models to understand and classify text accurately [3].

The application of domain-specific NLP models, such as scispaCy, has further improved the accuracy and relevance of text classification in specialised fields like biomedical research and clinical practice [4]. ScispaCy, a specialised extension of the spaCy NLP library, is tailored for scientific and clinical text, making it highly effective for tasks that require an understanding of domain-specific terminology and contexts [4,5].

A major breakthrough in NLP came with the development of transformer-based models, leading to the rapid rise of large language models (LLMs) [6]. Among these, Generative Pre-trained Transformer (GPT) models, such as ChatGPT, have gained widespread recognition for their autoregressive capabilities [7]. In contrast, Bidirectional Encoder Representations from Transformers (BERT) models, developed by Google, can understand the context of words in both directions, offering a different and often superior approach for NLP tasks [8]. These foundational models require extensive pre-training on large text corpora and subsequent fine-tuning on specific tasks using labelled datasets to achieve high performance in domains like text classification, named entity recognition, and span categorisation. Domain-specific adaptations, such as Bio-clinicalBERT for medical texts, further enhance their utility in specialised fields by using transfer learning to improve their applicability and accuracy in various biomedical NLP tasks [9–12].

Currently, the UK-based National Vascular Registry (NVR) gathers national data on primary AAA repairs, revision AAA repairs, lower limb bypass, lower limb angioplasty, major limb amputation, and carotid endarterectomy. The registry relies on the manual entry of patient data, which is not only time-consuming but also necessitates manual tracking for follow-up data such as outcomes at 30 days post-procedure. The adoption of NLP technologies could significantly reduce this burden by automating data extraction and analysis processes [1,13].

In vascular surgery, NLP applications have demonstrated significant potential. One such use has been the identification of AAA diagnoses from radiology reports streamlining the process for surveillance or referral initiation [14,15]. AAA features such as maximal diameter within the reports have also been extracted to guide the next management steps [16]. NLP has also shown the potential to be used as a decision support system for predicting aortic dissection, highlighting emergency department doctors for appropriate investigation and management with a reported AUC of 0.90 [17]. Beyond aortas, studies have demonstrated the use of NLP to more accurately identify patients with peripheral artery disease (PAD) from EHRs and radiology reports [18,19]. Further models have been developed to identify complications such as chronic limb threatening ischaemia [20]. For carotid stenosis, NLP has been used to categorise stenosis severity from imaging reports, achieving a positive predictive value of 99% for ultrasound and 96.5% for CTA and MRA [21]. However, there is no published literature of NLP models trained to idenitify patients who have undergone AAA repairs nor whether they have undergone a primary or revision repair. The differentiation between primary and revision AAA repairs is of clinical importance due to there being two separate forms for NVR data collection of those undergoing primary or revision AAA repairs.

This paper outlines a framework for the development and comparison of four NLP models: a fine-tuned BERT-base LLM, a fine-tuned biomedical domain specific BERT LLM (*Bio-clinicalBERT*), a biomedical SpaCY based model (*scispaCy*) and a *Bio-clinicalBERT/scispaCy* ensemble model to identify and classify abdominal aortic aneurysm (AAA) repair cases from unstructured free-text EHRs into categories of primary repair and revision repair.

## Methods

### Data Collection and Preparation

The publicly available MIMIC-IV-Note dataset was used, comprising of 331,794 deidentified discharge summaries from 146,815 patients admitted to the Beth Israel Deaconess Medical Center in Boston, MA, USA between 2008 and 2022 [22]. The data was unstructured, free text clinical notes containing discharge summaries detailing admission clerking, observations, hospital course, pertinent radiology reports, blood test results, and relevant discharge instructions. All records were stripped of protected health information in compliance with the Health Insurance Portability and Accountability Act (HIPAA) Safe Harbor provision.

To simulate a real-life clinical utility whilst maintaining data privacy, the dataset was pseudo-anonymised using the Python Faker library. Identifiable information placeholders, such as names, unit numbers, dates of birth, admission and discharge dates, and attending physician names, were replaced with realistic fake data. Anonymised names and dates in the clinical narrative were not synthetically generated and a deidentification placeholder remained in place.

### Multi-Tiered Classification Approach

The classification model used a structured multi-tiered approach using three classification tasks to achieve the aim of classifying AAA repairs. Task 1 was trained to identify vascular surgery related admissions. The model was then used to extract vascular-related admissions from the dataset using a probability threshold of 0.2 to maximise recall. This subset dataset was further annotated and then used to identify AAA repair cases (Task 2) which was then used to extract AAA repair records from the vascular dataset using a threshold of 0.5. A further model (Task 3) was trained to classify these AAA cases into primary and revision repairs. Task 3 also included a ‘Non-AAA’ classification to be able to appropriately categorise cases that had been misidentified in previous steps. In this paper the scispaCy model was used to extract the data for each task as demonstrated in Figure 1.

1. **Task 1 and Task 2**: Models were fine-tuned using annotated datasets with two labels:
  a. Task 1 – Vascular or Non-Vascular
  b. Task 2 – AAA repair or Non-AAA repair
2. **Task 3**: Model was fine-tuned using an annotated dataset with 3 labels. Non-AAA cases were included in this classification as previous models were not 100% accurate and Non-AAA cases had been included within the dataset.
  a. Task 3 – Primary AAA Repair or Revision AAA Repair or Non-AAA

**Fig. 1.**
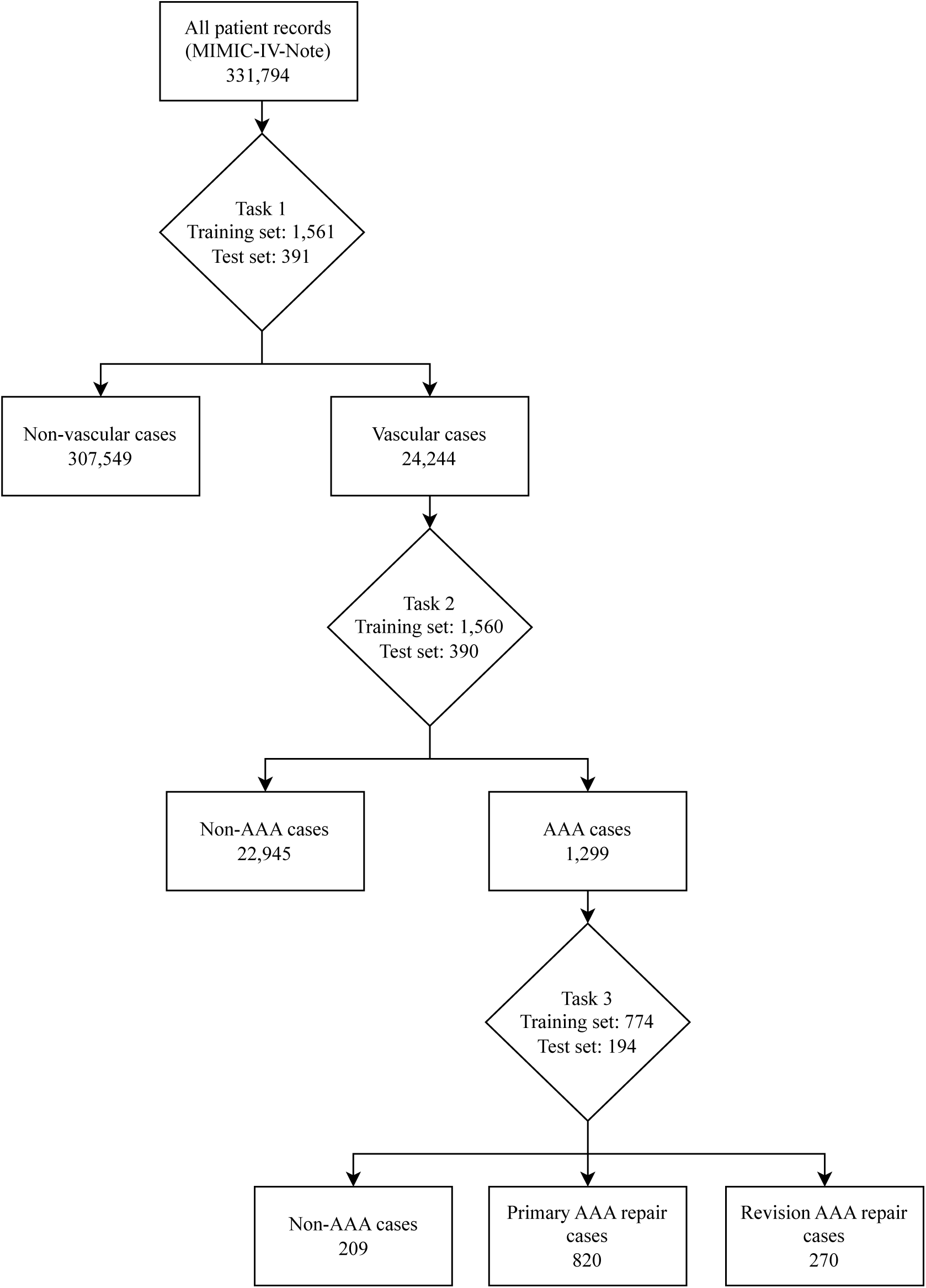
Flowchart showing the extraction of cases using a trained scispaCy model for each classification task. Task 1 – Vascular vs Non-Vascular classification performed using a threshold of 0.2. Task 2 – AAA vs Non-AAA classification performed using a threshold of 0.5. Task 3 – Primary AAA repair vs Revision AAA repair vs Non-AAA classification performed using a threshold of 0.5. AAA – Abdomional aortic aneurysm.

### Annotation

The data annotation process was conducted using Prodigy annotation software (Explosive AI, Berlin, Germany), by a Vascular Surgery Specialty Registrar. Terms related to vascular surgery and AAA repairs were used as ‘seeds’ for annotation, and an active learning approach was implemented utilising SciSpacy’s *en_core_sci_md model*. Seed terms can be found in appendix 1.

For Task 1, EHRs were categorised as ‘Vascular’ if there was a pathology relevant to vascular surgery during their admission as per National Health Service (NHS) England Service Specifications for Vascular Services [23]. Diabetic foot infections requiring debridement +/− revascularisation were also included. Stable chronic vascular conditions not being actively treated during the admission were annotated as ‘Non-vascular’. For Task 2, EHRs were categorised as ‘AAA repair’ if there was a repair of a thoracic and/or abdominal aortic and/or iliac aneurysm during the admission. Isolated ascending aortic aneurysm repairs were excluded. For Task 3, EHRs were categorised as ‘Primary AAA repair’ if a repair occurred on a previously untreated segment of the thoracic and/or abdominal aorta. ‘Revision AAA repair’ was defined as a further repair on a previously treated thoracic and/or abdominal aortic segment such as endoleak or aneurysmal disease at, or adjacent to, the anastomosis site.

A training curve function was utilised for each annotated dataset to determine the quality of the collected annotations, and whether more training examples would improve accuracy. Annotation was deemed sufficient if a training curve stopped showing improvement in the last 25% of the dataset. Annotated data was split into 80% for training and 20% for evaluation.

### SpaCy model training

The ‘*en_core_sci_md’* model from the open-source biomedical NLP library, scispaCy, using the spaCy Python framework, was used to develop an NLP pipeline [4,5]. The pipeline used ‘tok2vec’ tokenization and ‘textcat_multilabel’ components with other components frozen for training. The training process was configured with a dropout rate of 0.1, Adam optimizer with a learning rate of 0.001, and L2 weight decay. Training was performed for a maximum of 20 epochs using an Apple Macbook M2 Pro (California, USA) 16-core Graphics Processing Unit (GPU).

### BERT model training

#### Tokenization and Sliding Window Approach

Given the varying lengths of medical records, a sliding window tokenization approach was used to overcome the 512 maximum token sequence length constraint of BERT models. To accommodate BERT’s special tokens [CLS] and [SEP], the actual token window was set to 510 tokens. The BertTokenizer from the Hugging Face Transformers library was used for tokenizing the input texts. A stride of 255 tokens was used to allow sufficient overlap between segments, preserving contextual information across the segmented texts.

The segments were converted into model inputs, and probabilities for each segment were predicted using the BERT based models. Softmax function was applied to convert logits into probabilities, ensuring each class probability was normalised. Segment probabilities were then averaged across all segments of the record to obtain aggregated probabilities. A default threshold of 0.5 was applied to these aggregated probabilities to implement classification.

#### Model Architecture and Training

A *BERT-base-uncased* and a biomedical domain-specific BERT model (Bio-clinicalBERT) were used [8,9]. The fine-tuning process was conducted using a Google Colabatory (California, USA) Tensor Processing Unit (TPU) runtime environment using TensorFlow’s TPU strategy.

The models were compiled with the Adam optimizer. Sparse Categorical Crossentropy loss and accuracy were used as training evaluation metrics. Early stopping and learning rate reduction on plateau callbacks were implemented to prevent overfitting and dynamically adjust the learning rate.

The training process used a maximum of 5 epochs, with a batch size of 16. A validation set was used to monitor performance and adjust training dynamically through callbacks which included early stopping if validation loss did not improve for 2 consecutive epochs to reduce the chance of overfitting.

### Ensemble model

Predictions from both trained scispaCy and fine-tuned Bio-clinicalBERT models were integrated into a single pipeline. Predictions from the fine-tuned Bio-clinicalBERT model were obtained by averaging the softmax probabilities across all segmented texts. Predictions from the scispaCy model, configured with a multi-label text categorisation component (’textcat_multilabel’), were collected and processed. The aggregated probabilities from both models were then combined by averaging to form a combined prediction output.

### Evaluation Metrics

All inference was performed using a single Nvidia L4 (California, USA) GPU. SciSpaCy, BERT models and ensemble model were evaluated using accuracy, precision, recall and F1-score calculations per class using a probability threshold of 0.5. ROC curves were calculated with AUC values to assess class discrimination based on predicted probabilities.

## Results

A total of 4870 admission records were annotated with 1952 records annotated as vascular or non-vascular for Task 1, 1950 records annotated as AAA repair or non AAA repair for Task 2, and 968 records annotated as primary AAA, revision AAA repair or non-AAA repair for Task 3. Model training and inference times for BERT and scispaCy models are summarised in Table 1. The average training time across the 3 models was 3.03 mins/epoch for BERT-base, 2.75 mins/epoch for Bio-clinicalBERT and 2 mins/epoch for scispaCy. Inference time for scispaCy model was faster that BERT models with an average throughput of 2.87 samples/secs compared with 2.28 samples/secs for BERT-base and 2.32 samples/secs for Bio-clinicalBERT.

**Table 1:** Comparison of training and inference metrics for BERT and scispaCy models for Task 1 (Vascular vs Non-Vascular classification), Task 2 (AAA vs Non-AAA classification) and Task 3 (Primary AAA repair vs Revision AAA repair vs Non-AAA classification). Bio-clinicalBERT and BERT-base fine-tuned using Google Colab Tensor Processing Unit v2. ScispaCy trained using Apple Macbook M2 Pro 16-core Graphics Processing Unit (GPU). All inference performed using an Nvidia Tesla L4 GPU.

|  | Task 1 (n=1952) |  |  |  | Task 2 (n=1950) |  |  |  | Task 3 (n=968) |  |  |  |
| --- | --- | --- | --- | --- | --- | --- | --- | --- | --- | --- | --- | --- |
|  | BERT-base | Bio-clinical BERT | scispaCy | Ensemble | BERT-base | Bio-clinical BERT | scispaCy | Ensemble | BERT-base | Bio-clinical BERT | scispaCy | Ensemble |
| Annotated data for training | 1,561 | 1,561 | 1,561 | 1,561 | 1,560 | 1,560 | 1,560 | 1,560 | 774 | 774 | 774 | 774 |
| Annotated data for evaluation | 391 | 391 | 391 | 391 | 390 | 390 | 390 | 390 | 194 | 194 | 194 | 194 |
| Number of epochs for training | 4 | 3 | 20 | - | 4 | 4 | 20 | - | 3 | 3 | 20 | - |
| Fine-Tuning Time (mins per epoch) | 3.5 | 2.66 | 2 | - | 3.25 | 3.25 | 2 | - | 2.33 | 2.33 | 2 | - |
| Inference Time (Total, secs) | 186 | 199 | 153 | 347 | 181 | 163 | 133 | 296 | 75 | 74 | 62 | 136 |
| Inference Time (secs/sample) | 0.476 | 0.509 | 0.391 | 0.887 | 0.464 | 0.418 | 0.341 | 0.759 | 0.387 | 0.381 | 0.320 | 0.701 |
| Throughput (samples/secs) | 2.10 | 1.96 | 2.56 | 1.13 | 2.15 | 2.39 | 2.93 | 1.32 | 2.59 | 2.62 | 3.13 | 1.43 |

For Task 1, the scispaCy and ensemble models had the highest overall accuracy of 0.97 and BERT-base had the lowest accuracy of 0.92 (Table 2). Bio-clinicalBERT demonstrated the worst recall for classification of vascular cases of all models with 0.70 compared to 0.79 for BERT-base and 0.89 for scispaCy and ensemble models. ScispaCy had the best discriminative ability with an AUC of 0.99 and Bio-clinicalBERT and BERTbase had the worst discrimination with 0.95 (Figure 2).

**Fig. 2.**
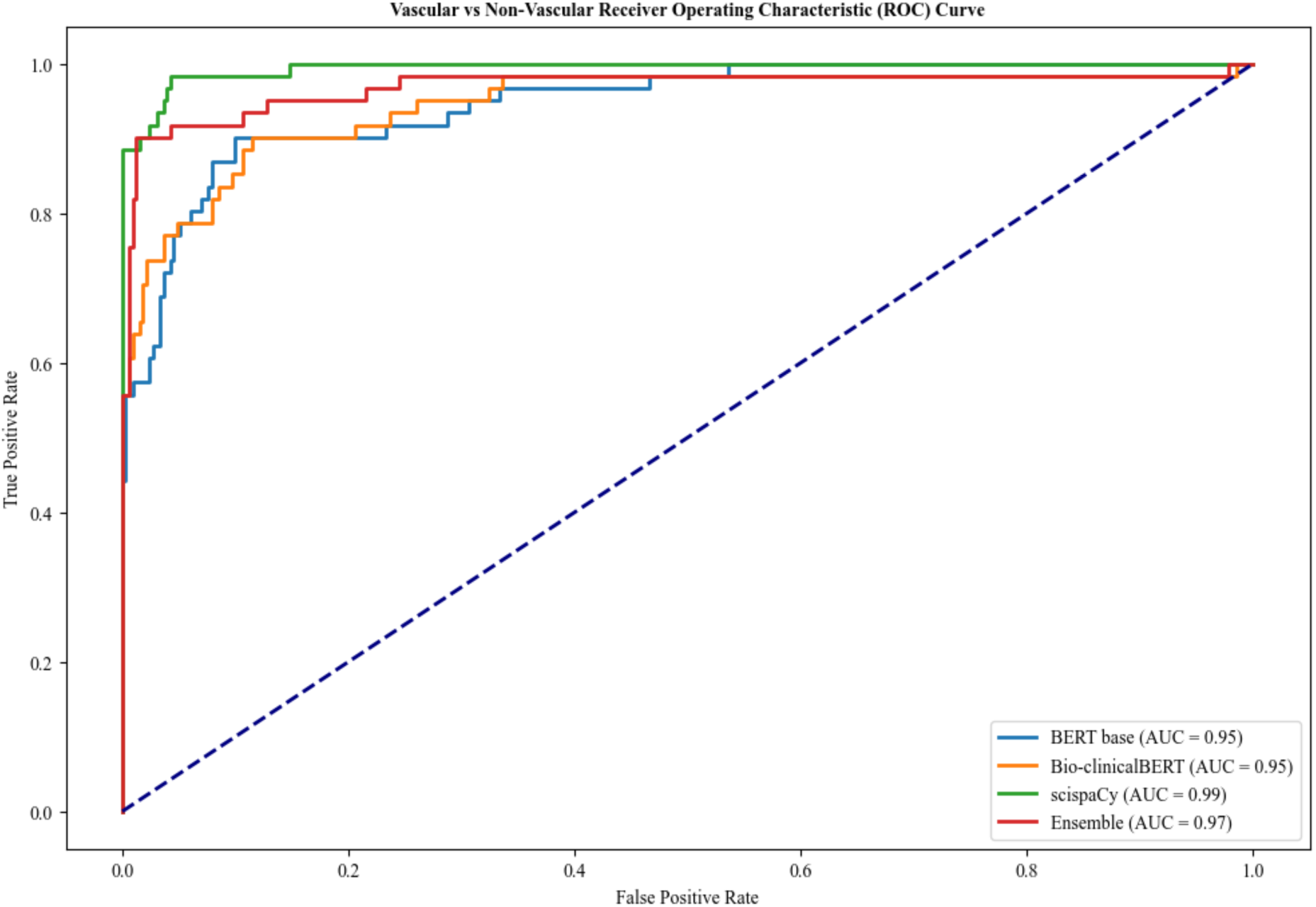
Comparison of receiver operating characteristic curves for Task 1 – Vascular vs Non-Vascular classification.

**Table 2:** Table showing classification report for Task 1 – classifying admissions for patient with acute vascular conditions with a threshold of 0.5.

| Model | Accuracy | Vascular |  |  | Non-Vascular |  |  |
| --- | --- | --- | --- | --- | --- | --- | --- |
|  |  | Precision (Vascular) | Recall (Vascular) | F1-Score (Vascular) | Precision (Non-Vascular) | Recall (Non-Vascular) | F1-Score (Non-Vascular) |
| BERT-base | 0.92 | 0.74 | 0.79 | 0.76 | 0.96 | 0.95 | 0.95 |
| Bio-clinicalBERT | 0.94 | 0.88 | 0.70 | 0.78 | 0.95 | 0.98 | 0.96 |
| ScispaCy | 0.97 | 0.92 | 0.89 | 0.90 | 0.98 | 0.98 | 0.98 |
| Ensemble | 0.97 | 0.93 | 0.89 | 0.91 | 0.98 | 0.99 | 0.98 |

For stage 2, all model performed very well across all metrics as shown in Table 3. The ensemble, scispaCy and Bio-clinicalBERT models showed the highest degree of accuracy of 0.99 with all models achieving an AUC of 1.00 (Figure 3). BERTbase had a slightly worse recall of 0.97 when compared with the other models for AAA classification (0.98 and 1.00).

**Fig. 3.**
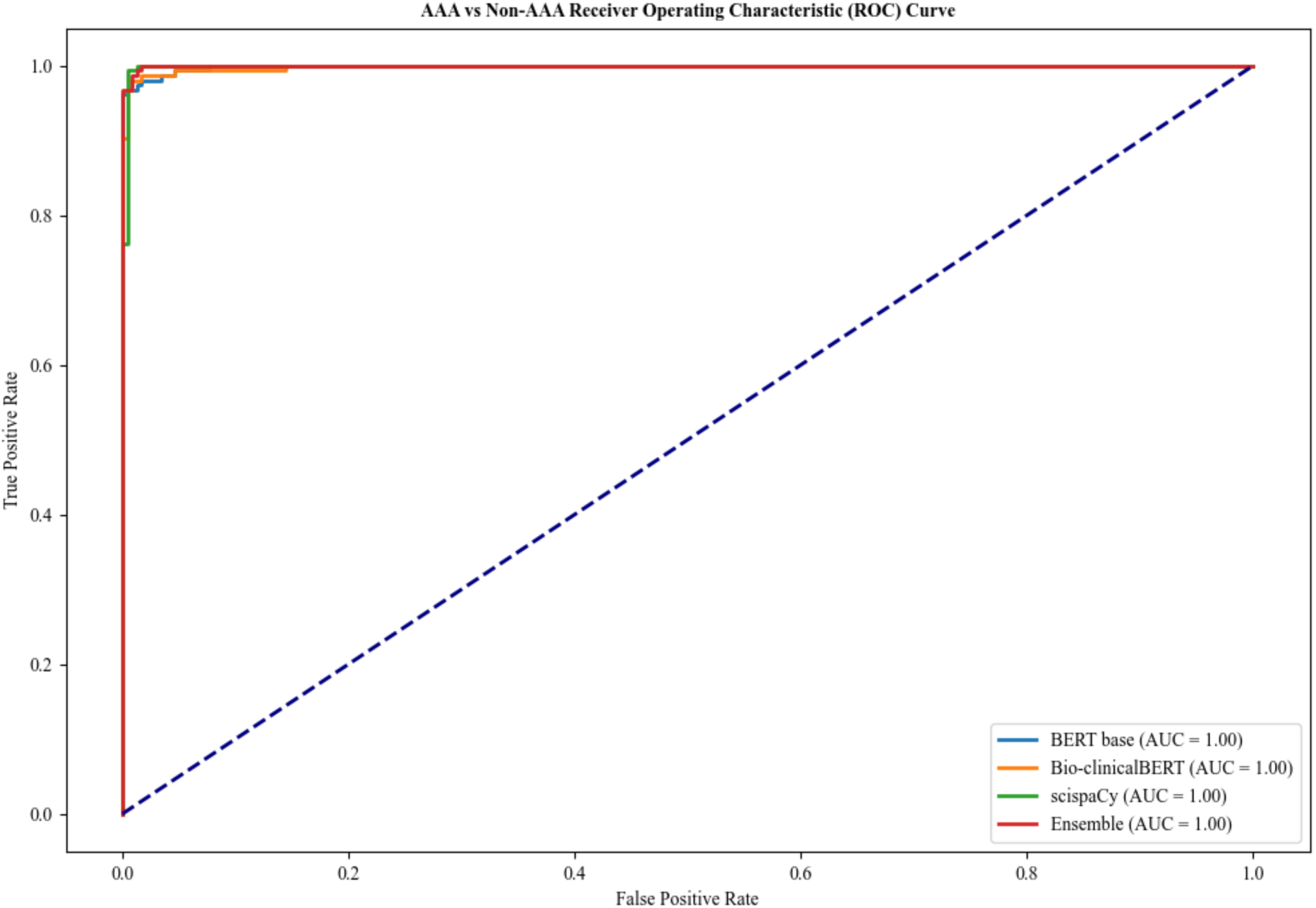
Comparison of receiver operating characteristic curves for Task 2 – AAA vs Non-AAA classification.

**Table 3:** Table showing classification report for stage 2 – classifying admissions for patients who undergone AAA repair during their admission with a threshold of 0.5.

| Model | Accuracy | Non-AAA |  |  | Primary AAA repair |  |  | Revision AAA repair |  |  |
| --- | --- | --- | --- | --- | --- | --- | --- | --- | --- | --- |
|  |  | Precision | Recall | F1-Score | Precision | Recall | F1-Score | Precision | Recall | F1-Score |
| BERT-base | 0.92 | 0.97 | 0.96 | 0.96 | 0.90 | 0.95 | 0.93 | 0.86 | 0.72 | 0.78 |
| Bio-clinicalBERT | 0.98 | 1.00 | 0.96 | 0.98 | 0.97 | 0.99 | 0.98 | 0.96 | 1.00 | 0.98 |
| ScispaCy | 0.93 | 0.99 | 0.93 | 0.96 | 0.93 | 0.93 | 0.93 | 0.79 | 0.92 | 0.85 |
| Ensemble | 0.93 | 0.99 | 0.93 | 0.96 | 0.93 | 0.94 | 0.94 | 0.82 | 0.92 | 0.87 |

For Task 3, Bio-clinicalBERT and ensemble models achieved an AUC of 1.00 (Figure 4) with Bio-clinicalBERT displaying the best overall accuracy of 0.98 (Table 4). Bert-base had the lowest F1-score across all classification tasks for Task 3 with Bio-ClincialBERT having the highest.

**Fig. 4.**
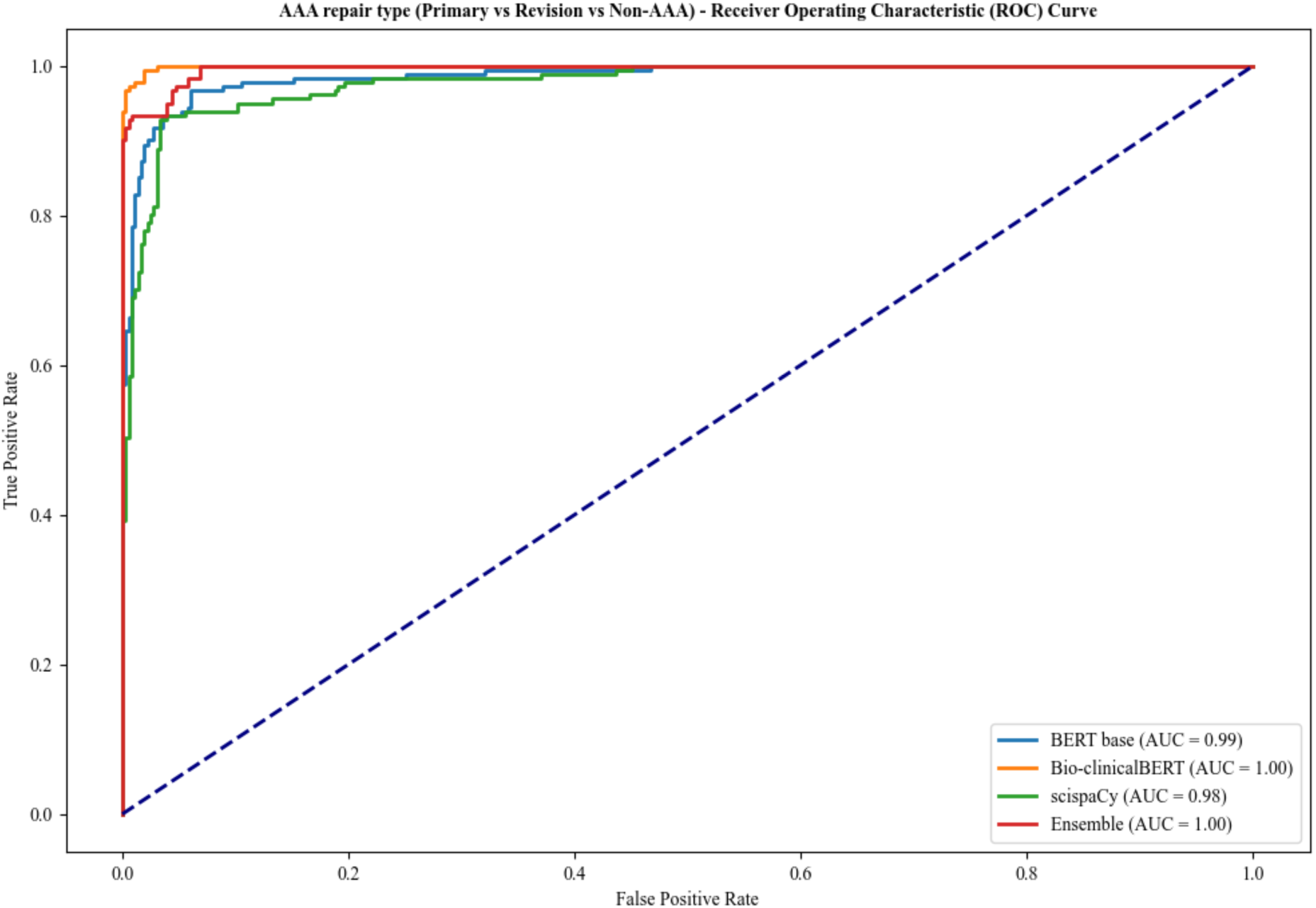
Comparison of receiver operating characteristic curves for Task 3 – Primary AAA repair vs Revision AAA repair vs Non-AAA classification.

**Table 4:**
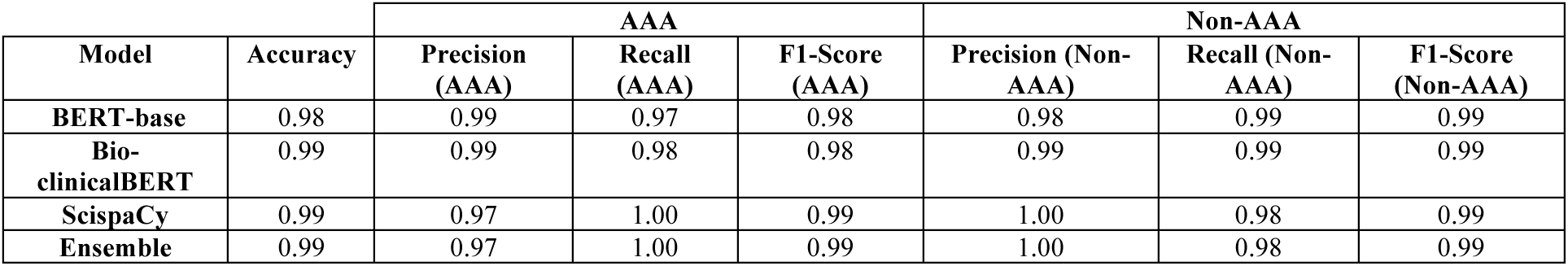
Table showing classification report for Task 3 – classifying type of AAA repair undertaken during admission with a threshold of 0.5.

Across the three classification tasks in this paper Task 2 showed the most consistent highly accurate results across all models. The ensemble model consistently had the highest or joint highest f1-score across all classification tasks, apart from Task 3, demonstrating its robustness and reliability. BERT-base was the worst performing model across all tasks with the lowest f1-scores.

## Discussion

This multi-tiered NLP approach demonstrates a proof of concept for tackling classification tasks in healthcare, shown here by the identification and classification of AAA repairs from unstructured EHRs. The tiered approach allows for iterative refinement of the dataset, starting with broad classifications and progressively narrowing down to more specific categories. This method could be particularly useful in scenarios where traditional categorisation methods, such as ICD-10 codes, lack sufficient granularity for certain conditions, like acute limb ischemia [24]. The integration of NLP models into existing EHR infrastructure has the potential to enhance data extraction and auditing processes across various medical specialties and other domains dealing with unstructured text data.

While several studies have explored the use of NLP in cardiovascular medicine, to the best of our knowledge, this is the first study that specifically focuses on using NLP models to identify patients who have undergone an AAA repair during their admission and to classify the repair as either primary or revision [14–16,25]. The most closely related work is by Weissler et al., who developed an NLP model called ‘PAD-ML’ to identify patients with peripheral arterial disease (PAD) from EHRs using an artificial neural network approach [18]. PAD-ML achieved an AUC of 0.888. In contrast, our overall best-performing model, the scispaCy/Bio-clinicalBERT ensemble, achieved AUROCs of 0.99, 1.00, and 1.00 for identifying vascular patients, AAA repairs, and classifying repairs as primary or revision, respectively. Although both Bio-clinicalBERT and scispaCy performed well individually, combining these methods resulted in more robust performance across all tasks. This highlights the effectiveness of integrating domain-specific embeddings, contextual understanding, and state-of-the-art NLP architectures for clinical tasks. While direct comparisons with PAD-ML are limited due to differences in tasks, datasets, and metrics, our models demonstrate strong results in the context of related work [14–16].

The comparison between BERT-base, Bio-ClinicalBERT, and scispaCy highlights the trade-off between model complexity and performance in clinical NLP tasks. The domain-specific Bio-ClinicalBERT outperformed BERT-base, particularly in distinguishing primary and revision AAA repairs showing the effectiveness of transfer learning in LLMs. However, the lighter-weight scispaCy model achieved comparable performance while being more computationally efficient. With the development of increasingly complex generalised LLM models such as GPT-4 and Llama 3, and larger clinical foundation models such as GatorTron, clinical NLP developer will need to balance model complexity, performance and interpretability to ensure that the model delivers on accuracy whilst maintaining computational efficiency and transparency in decision-making [26].

There are limitations to the models developed in this paper which need to be considered in future development and implementation. The model was annotated by a single clinically trained annotator, which whilst following a structured annotating guide, reduces the reliability of the annotated dataset. Future models intended for clinical use would require multiple clinically trained annotators to ensure reliability of the training dataset. The models were trained on a dataset from one hospital in the USA, thereby narrowing the breadth of medical language and style used in medical record keeping across the world. There were some differences in vascular practices identified in the US medical records compared to standard UK NHS practice, such as the management of acute diabetic foot infections by podiatry in the US EHRs. Although fundamental clinical practice is similar across countries, there may be subtleties in the records which could reduce the generalisability of the models when used on different datasets. External validation of the models would assist in identification of this issue. The methodology outlined in this paper provides a framework for fine-tuning/training of both LLMs and NLP models enabling models to be trained and validated locally, thereby capturing local variation in practices and not relying on the generalisability of an externally trained model.

Future work should include validating the model across multiple healthcare institutions to assess its generalisability and robustness. This can help identify any institution-specific biases and need for model refinement. The integration of other NLP tasks such as named entity recognition and/or span recognition would allow vascular pathology specific data (e.g AAA diameter) to be identified and extracted from unstructured free text data, potentially enabling the automated data extraction and upload to national registries such as the NVR.[16] The development of such a pipeline would remove barriers such as administrative and time burden to allow a much broader data capture of patients. Additionally, integrating other data modalities such as imaging would allow AAA morphology to be captured in detail helping with predictive modelling [27]. The pipeline should also incorporate a clinician-in-the-loop mechanism that allows clinicians to provide feedback to the model and contribute to ongoing model refinement. Other than using the framework developed in this paper to identify other NVR-relevant patients such as lower limb bypass or carotid endarterectomies the technique could be expanded to other medical speciality registries such as National Joint Registry or National Confidential Enquiry into Patient Outcome and Death.

This study presents a robust framework for developing NLP models to identify and classify AAA repairs from unstructured EHRs. The high accuracy achieved by our models demonstrates the potential for implementing these techniques in clinical data pipelines to improve auditing of AAA repairs at both local and national levels. Our work represents a significant step towards harnessing the untapped potential of unstructured EHR data across various medical specialties. By automating the extraction and classification of pertinent clinical information, NLP models can reduce administrative burden, increase data capture, and ultimately contribute to improved patient care and outcomes in vascular surgery and beyond.

## Statements and Declarations

Funding: This research received no grants from any funding source

Conflict of interest statement: No authors of this manuscript have any declared conflicts of interest

Data/code availability: Models are available for download and use under GNU General Public License v3.0 at https://huggingface.co/danielcthompson with demonstration scripts available at https://github.com/dc-thompson/AAA_classification. The MIMIC IV dataset used for training is available at https://physionet.org/content/mimic-iv-note/2.2/. The underlying code and annotated dataset for model training used in this study is not publicly available but may be made available to qualified researchers on reasonable request to the corresponding author.

## Appendix 1

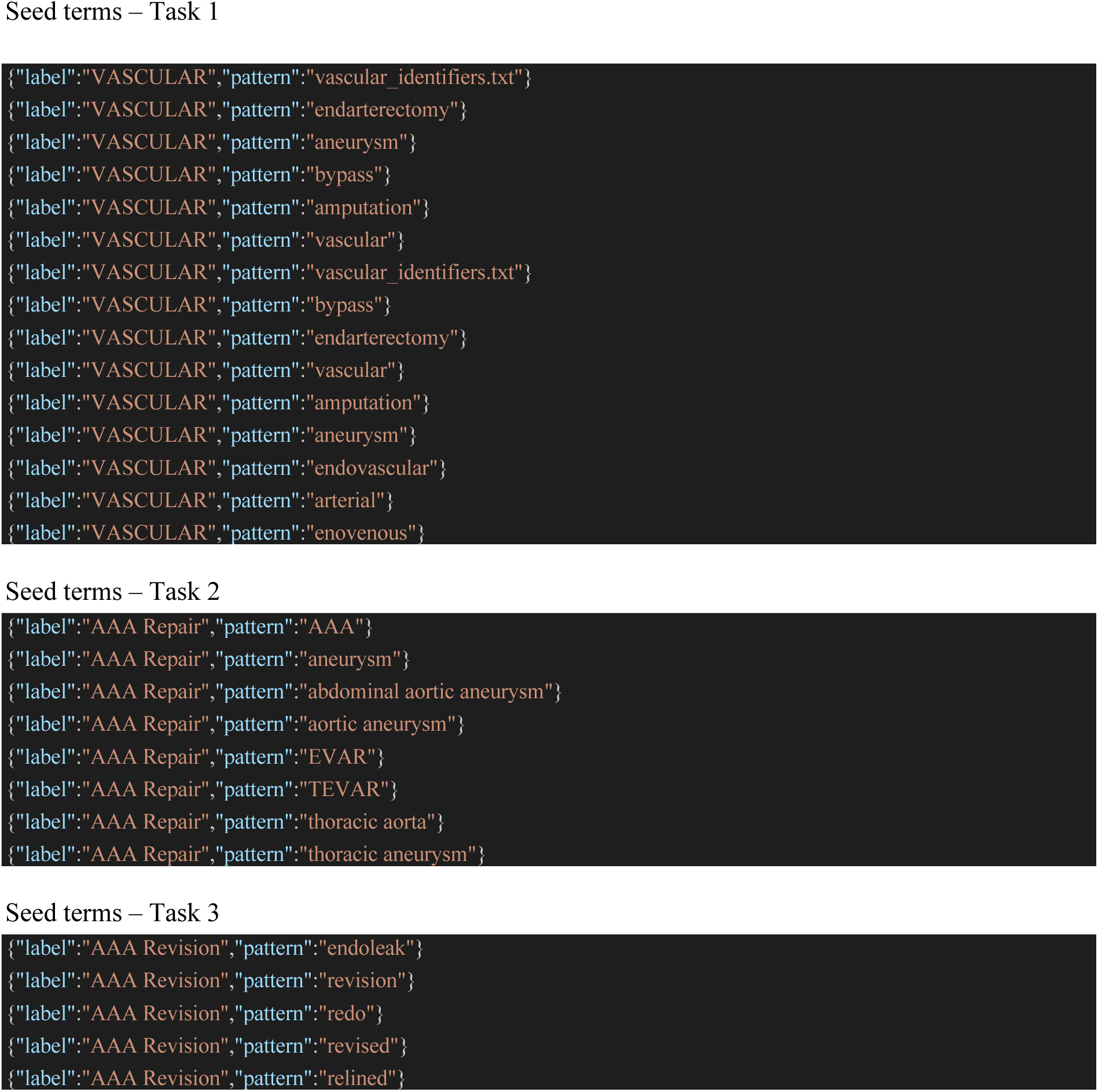

